# Accuracy and Precision of Multiple Laboratory and Field Methods to The Criterion *In Vivo* Five-Compartment Body Composition Model and Their Association with Muscle Strength in Collegiate Athletes of Varying States of Hydration: The *Da Kine* Protocol Study

**DOI:** 10.1101/2023.05.30.23290630

**Authors:** Devon Cataldi, Jonathan P. Bennett, Michael C. Wong, Brandon K. Quon, Yong En Liu, Nisa Kelly, Thomas Kelly, Dale A. Schoeller, Steven B. Heymsfield, John A. Shepherd

## Abstract

**Objective:** To compare multiple body composition analysis methods in athletes with varying states of hydration to the criterion 5-compartment model(5CM) of body composition and assess the relationships of technique-specific estimates of fat and fat-free mass(FM, FFM) to muscle strength.

**Methods:** Body composition was assessed in 80(40-female) athletes with a mean age of 21.8±4.2 years. All athletes underwent laboratory-based methods: air-displacement plethysmography(ADP), deuterium-oxide dilution(D_2_O), dual-energy X-ray absorptiometry(DXA), underwater-weighing(UWW), and field-based: 3D-optical(3DO) imaging, and three bioelectrical impedance(BIA) devices(S10/SFB7/SOZO). Participants’ muscular strength was assessed by isokinetic/isometric dynamometry. Accuracy was assessed by Lin’s concordance correlation coefficient(CCC) and precision by root-mean-square coefficient of variation(RMS-CV%).

**Results:** Athletes’ hydration status(total body water/FFM) was significantly(*p*<0.05) outside of the normal range in both males(0.63-0.73%) and females(0.58-0.78%). The most accurate techniques(ADP/DXA) showed moderate-substantial agreement(CCC=0.90-0.95) in FM and FFM, whereas all field assessments had poor agreement(CCC<0.90), except 3DO FFM in females(CCC=0.91). All measures of FFM produced excellent <1.0% precision, whereas FM from ADP, DXA, D_2_O, S10, and UWW had <2.0%. The associations between muscle strength and the various devices’ FFM estimates differed. However, more accurate body composition compared to the criterion produced a better determination of muscle strength by significant quartile *p*-trends(*p<*0.001). The 5CM exhibits the highest determination for all categories of muscle strength which persisted across all hydration measures.

**Conclusion:** To optimize accuracy in assessing body composition and muscle strength, researchers and clinicians should prioritize selecting devices based on their accuracy compared to the 5CM. Reliable approaches such as ADP and DXA yield accurate and precise body composition estimates and thereby, better strength assessments, regardless of hydration status. Future athlete studies should investigate the impact of changes in FFM on functional measures compared to the criterion method.

**Summary Box:** This study compared various body composition analysis methods in athletes with varying states of hydration to the criterion 5-compartment model(5CM) and assessed their relationship to muscle strength. The results showed that accurate and precise estimates of body composition can be determined in athletes, and a more accurate body composition measurement produced better strength estimates. The best laboratory-based techniques were air displacement plethysmography(ADP) and dual-energy x-ray absorptiometry(DXA), while field assessments had moderate-poor agreement. Prioritize accurate body composition assessment devices compared to the 5CM for better strength estimates in athletes.

## INTRODUCTION

Fat-free mass(FFM) is a functional, metabolically active tissue that contributes to strength and force production and plays a key role in sports performance^1^. When fat mass(FM) is in excess, it can hinder performance and adversely affect physiological systems, such as the endocrine system by increasing the production of cortisol and leptin, which play crucial roles in the nervous system for thermoregulation, as well as the immune system through heightened inflammation, all of which collectively impact athletic performance^2^. Several studies have reported moderate correlations between FM and BMI when gender and age are considered^3, 4^. However, strong evidence suggests that athletic body types play a critical role in the relationship between FM and BMI, leading athletes, clinicians, and coaches to shift away from solely using BMI to assess performance^5^.

To adhere to sports criteria, many athletes use extreme weight-control methods that can be detrimental to both health and performance due to continuous dieting, energy deficits, and/or extreme weight-loss practices^6^. As such, errors in body composition assessments can lead to improper conclusions on athletic programming, impacting health and performance^7^. Accurate body composition assessments can also help identify and monitor relative energy deficiency(RED-S) risk and other injuries or illnesses^8^, as well as enable athletes to adjust their training and nutritional habits to the demands of their sport^9^.

The relationship between body composition and muscle strength is unclear due to FFM’s complex physiological composition(i.e. water, protein, minerals, and others) between participants and populations, and potentially how the estimate is determined^10^. An ideal body composition method would be accurate/precise and provide the highest muscle strength/performance associations. Multiple studies have explored absolute or relative proportions of body composition and their associations with muscle strength, though not comprehensively comparing multiple systems, muscle groups, or other predictive factors in athletes of varying states of hydration, as defined by total body water(TBW)/FFM. Furthermore, Nickerson and Gudivaka emphasized the importance of considering skin hydration/temperature status in athletic populations to obtain accurate measurements of body composition estimates^11,12.^

The reference method to assess body composition *in vivo* is the 5-compartment model(5CM), which combines criterion assessments for each of the individual components of TBW by deuterium-oxide dilution(D_2_O), bone mineral content(BMC) by dual-energy X-ray absorptiometry(DXA), total body volume(BV) by air displacement plethysmography(ADP) or under-water weighing(UWW), and body mas(BM) to accurately quantify whole-body FM^13^. Despite the well-known strengths of multicompartment modeling, it is often limited by the time and expense necessary to produce them^14^. Few studies have examined a variety of clinical body composition assessment techniques to the 5CM in athletics, where the sample sizes have been small and device inclusion is limited^15, 16^. Additionally, rapid field techniques such as bioelectrical impedance analysis(BIA) and 3D-optical imaging(3DO) are becoming more accessible in athletic training facilities^15, 17-19^. In performance assessment studies, accuracy is crucial, but the relationship between body composition measures and sport-specific performance outcomes like muscle strength in athletes is not fully understood.

Therefore, this study aims to examine the accuracy of different laboratory and field-based body composition methods to a criterion model and show their associations to muscle strength within a collegiate athlete population. Additionally, we explored the impact of skin hydration and temperature variations to create a more accurate 5CM to improve strength predictions. We hypothesized that field-based methods, when evaluated, would offer similar accuracy and precision of body composition estimates in their associations to muscle strength over the laboratory-based methods.

## METHODS

### Experimental design

The *Da Kine* Study is a cross-sectional observational study of athletes to examine the association of body composition estimates to muscle strength. This study was approved by the University of Hawai’i Research Compliance and Institutional Review Board(IRB), protocol#2018-01102. Participants provided written consent.

### Participate Involvement

The patient-centered *Da Kine* study involved patients in the research process. Patient input was obtained through focus group sessions and interviews to shape the research question, outcome measures, and recruitment methods. A patient joined the trial steering committee. The University of Hawaii Clinical Trials Department prioritized the study for cancer clinicians. Participants received trial results via email and could access updates through a dedicated website(https://shepherdresearchlab.org/) and a non-specialist study newsletter.

### Participants

Between April2019-March2020, eighty healthy male and female collegiate and intramural athletes(>18years) representing various BMI ranges were enrolled. Athletes were recruited during their in-season or off-season strength and conditioning routines, with investigators approaching coaches and trainers during practice. Exclusions included pregnancy/breastfeeding, metal implants, or recent body composition-altering procedures. Participants fasted and abstained from alcohol for at least eight hours before testing, and avoided moderate-intense exercise for 24hours. On the testing day, participants arrived at the University of Hawaii Cancer Center, adhered to pretesting protocols, and underwent anthropometry, thigh, and trunk strength tests, and height and weight measurements on a stadiometer(Seca264, Chino, CA). Ethnicity was self-reported.

**Supplemental Table 1** shows all methods obtained and an encompassing comparison of devices and their assumptions. All methods were taken in duplicate to calculate precision.

#### Laboratory-based methods

### Air Displacement Plethysmography

Measurements were taken using ADP in a BodPod(v5.4.1, COSMED, Concord, CA) to providing BV measurements required for 5CM, along with the standard output of body composition. Measurements were taken via the manufacturer’s standard protocol, where participants dressed in form-fitting attire, with a hair cap. The BodPod measures BV with corrections for residual lung volume and surface area artifacts(SAA)^20^. Thoracic gas volume(TGV) was measured by breathing through a tube connected to a filter and reference chamber, following the manufacturer’s instructions, or estimated if participants could not obtain a valid measurement directly due to the inability to achieve consistency over the three repeated TGV measurements(n=26). The BodPod software automatically calculated the SAA. These two adjustments(TGV and SAA) were factored into the overall BV calculation.

### Dual-energy X-ray absorptiometry

Whole-body DXA scans were performed using a Hologic Discovery/A system(Hologic, Marlborough, MA) to provide BMC to calculate osseous mineral(Mo) for 5CM, along with standard outputs of body composition. The scans were analyzed by a trained technologist using Hologic Apex version 4.5. Whole-Body Fan Beam and the National Health and Nutrition Examination Survey Body Composition Analysis(NHANES BCA) calibration option were disabled. DXA systems were calibrated according to standard Hologic procedures and all scans were taken by standard procedures^21^. DXA-derived BV was determined by Wilson et al in 2013^22^.

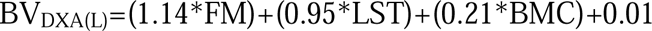

### Deuterium Oxide Dilution

TBW(for 5CM), FM, and FFM were determined using the D_2_O protocol defined in the International Atomic Energy Agency(IAEA) standards^23, 24^. A high-precision scale was used for D_2_O dosing(Denver Instrument M-310). All study participants provided the required two post-dose saliva samples. Based on previous research using multiple samples and technologies, saliva was chosen as the criterion^14, 25^. The saliva data was interrogated with a quality control method of a 5% difference between time points of three and four hours. If the difference was higher than 5%, the saliva samples were deemed to not have reached equilibrium and were excluded. Participants were provided with a measured dose of 30g(99.9%pure) D_2_O(Cambridge Isotope Laboratories, Tewksbury, MA) and 100ml local drinking water as a rinse to ensure the entire dose was consumed. During the four-hour D_2_O equilibration period, participants were allowed to consume up to 500mL of water which was recorded.

### Underwater Weighting

UWW measured BV and estimated body composition. On land, participants were weighed in their form-fitting suit caps, then entered the temperature-stable water with a nose clip. Immersed weight was measured using an electronic weighing system(EXERTECH, Dresbach, MN), transmitting data to a computer and providing continuous recording. Trials were performed at residual volume after maximal expiration. Participants sat on the UWW scale, slowly submerged, fully exhaled, and remained still for underwater weight measurement. This procedure was repeated three times and averaged.

### Bioelectrical Impedance Analysis/ Spectroscopy

Body composition was estimated in participants by three different systems: SOZO(ImpediMed, Carlsbad, CA), SFB7(ImpediMed, Carlsbad, CA), and S10(MF-BIA; InBody, Cerritos, CA). Each method was performed as per their respective device manufactural recommendations. The S10 and SFB7 scans were performed in an order of convenience approaching a random order immediately following DXA to allow for proper fluid normalization in the supine position^26^. Before each scan, participants cleaned their ankles, hands, and feet with alcohol wipes. For the SFB7 system, participants were tested using single-tab adhesive electrodes after lying supine for 10 minutes.

### Three Dimensional Optical Scans

Each participant underwent 3DO whole-body surface scans, with repositioning, on a Fit3D Proscanner with software version 4.1(Fit3D, Inc., Redwood City, CA). The 3DO scanner provided BV, FM, and FFM for analysis. The 3DO scanner is comprised of light-coding depth sensors, a rotating platform, and analysis software^19^. Participants stood on the turntable with legs separated and arms extended and holding the positioning handles following the protocol from the manufacturer. During the scans, the platform rotates 360 degrees over a period of 30-40 seconds, with the camera system emitting light and reflections being recorded by the camera.

### Skin Moisture and Temperature

A moisture meter(Moisture-Meter-D, Delfin Technologies) assessed tissue hydration using a control unit transmitting a 300MHz signal to a skin probe, functioning as an open-ended co-axial transmission line^27, 28^. The reflected wave depended on tissue dielectric constant, shown on the unit(range:1-80, pure water ≈80). Medium probes assessed tissue water at 1.5mm depth. Skin temperature was recorded using an infrared temperature scanner(Dermatemp DT-1001, Exergen, Newton, MA) before BIA scan. After BIA assessments and 10 minutes of supine position, both measurements were taken twice at three sites on the right side(forehead, dorsal hand, foot).

### Strength Assessments

Whole-body muscle strength was evaluated using an isokinetic dynamometer(Humac NORM, Computer Sports Medicine, Stoughton, MA). Participants were positioned at 95° trunk-to-thigh angle and secured with straps to stabilize their lower leg, thigh, and waist. They underwent warm-up, practice, and then performed five isometric and concentric repetitions of knee extension/flexion. After resting, they completed five maximal effort repetitions of trunk flexion/extension, followed by 15 consecutive repetitions. Data collection followed the Humac NORM manual^29^ protocol without gravity correction. Participants were instructed to exert maximum force rapidly, receiving verbal encouragement but without real-time feedback. The primary measure of strength was isokinetic leg and trunk extension, which maximized whole-body muscle mass assessment, involving multiple muscles and producing a mean peak force of Nm.

### Multicompartment Body Composition Models

5CM body composition model described by Wang^30^ was used as our criterion method. The 5CM includes BV by ADP, TBW and soft tissue mineral(M_s_) by D_2_O,(M_o_) by DXA by DXA, and BM, as shown in

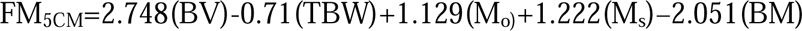

where M_o_ was calculated by total body BMC*1.0436, and M_s_ was calculated by TBW*0.0129^31,32^. For reporting 5CM_FFM_, BM was subtracted from 5CM_FM_ as outlined in For clarity, all other methods from each device will be that of FFM(with BMC) and not lean soft tissue(LST; without BMC)^33^. FFM was calculated by subtracting FM from BM, while density of FFM, total body density(Db), and hydration were calculated using the following equation. These values were calculated to compare to the standard reference values^34^.

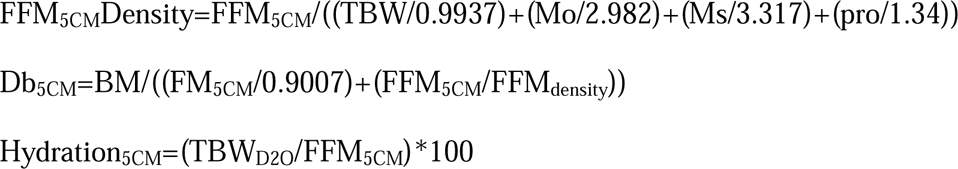

### Statistical Analysis

Shapiro–Wilks’s test was used to test normality and analysis of variance with post-hoc comparisons of participants with available data on all devices/testing methods to assess for significant mean differences. Agreement between FM and FFM errors between technologies and the 5CM were calculated using the root mean square error(RMSE), coefficient of determination(R^2^), intercept values, and Lin’s concordance correlation coefficient(CCC). The CCC agreement cutoffs are defined as follows: poor(<0.90), moderate(0.90-0.95), substantial(0.95-0.99), and almost perfect(>0.99)^35^. Precision was calculated as root-mean-square coefficient of variation(RMS-CV%). Stepwise linear regression was used for the predictor variable methods of BIA-TBW, skin temperature, and moisture to the outcome of TBW and strength estimates by using a *p*<0.10 to enter the model *p*<0.05 to stay in the model. Bootstrapping(n=1000) 95% confidence intervals for the R^2^ of each model using the percentile method were used to compare model performance. Additionally, Pearson’s correlation was performed for each body composition method to strength. All statistical calculations were performed using SAS 9.4(SAS, Cary, NC) and are consistent with the CHAMP-statement^36^.

## Results

In this study, 70 participants(35females) were included in the final analysis, where **Supplemental Figure 1** provides details of the data that was included and excluded. Due to a malfunction of the UWW device during data collection, only 24 participants(14 females) completed the test. Necessitating the use of separate matched 5CM comparisons for all analyses relating to the UWW device. Descriptive statistics are found in **Table 1** and determined to be normally distributed, including the strength measures. Males’ BMI ranged from 20.2-32.8 and 17.8-30.9 in females, whereas the FM ranged from 3.2-22.9kg in males to 3.4-26.4kg in females. Compared to Brozek1963 reference body both sexes in the current study demonstrated significant differences(*p*<0.05) in density and percentage values.

**Table 1.** Descriptive Statistics of Demographics, Whole Body Composition and Muscle Strength in Male and Female Athletes

|  | Variable | Units | Male (N=35) |  | Female (N=35) |  |
| --- | --- | --- | --- | --- | --- | --- |
|  |  |  | Mean (SD) | Min (Max) | Mean (SD) | Min (Max) |
| Demographics | Weight | kg | *82.27±10.09 | 61.9-102.5 | 62.97±10.44 | 43.9-92.4 |
|  | Height | cm | *180.99±10.26 | 159.3-203 | 168.04±8.86 | 154.7-188.1 |
|  | Age | years | 24.43±5.11 | 18-37 | 21.86±4.19 | 18-35 |
|  | BMI | kg/m<sup>2</sup> | *25.2±3.21 | 20.15-32.79 | 22.25±2.89 | 17.79-30.94 |
| Skin | Temperature | deg. | 34.17±0.83 | 32.5-35.6 | 33.97±1.14 | 31.1-35.6 |
|  | Moisture | F/m | *42.17±7.75 | 31-53.4 | 37.16±4.84 | 27.6-46.1 |
| ISOK Strength | LEG Ext | Nm | *139.08±34.44 | 67-200 | 94.11±27.94 | 54-155 |
|  | TRK Ext | Nm | *188.14±61.68 | 75-306 | 93.09±38.2 | 40-214 |
| Whole Body | ADP BV | L | *77.42±10.16 | 57.31-96.94 | 60.11±10.44 | 40.26-88.27 |
|  | D<sub>2</sub>O TBW | L | *49.94±6.42 | 36.3-60 | 34.14±6.01 | 22.8-49.03 |
|  | BMC | kg | *3.09±0.38 | 2.38-3.91 | 2.33±0.4 | 1.7-3.53 |
| 5C Model | 5C FM | kg | 11.58±5.9 | 3.18-22.87 | 14.36±5.66 | 3.39-26.36 |
|  | 5C FFM | kg | *71.25±8.53 | 54.79-84.33 | 48.87±7.92 | 33.87-69.48 |
|  | Mo | kg | *3.23±0.39 | 2.48-4.08 | 2.44±0.41 | 1.78-3.68 |
|  | Ms | kg | *0.64±0.08 | 0.47-0.77 | 0.44±0.08 | 0.29-0.63 |
|  | Protein | kg | *17.43±2.35 | 13.03-22.41 | 11.86±2.1 | 7.61-16.14 |
|  | FFM Density | g/cm<sup>3</sup> | †1.104±0.007 | 1.094-1.129 | †1.107±0.01 | 1.081-1.15 |
|  | TBW/FFM | % | †70.04±2.08 | 62.69-73.16 | †69.75±2.92 | 58.22-78.12 |
|  | Pro/FFM | % | †*24.51±2.11 | 21.52-31.6 | †24.35±2.88 | 16.09-35.03 |
|  | BMC/FFM | % | †4.36±0.37 | 3.61-5.31 | †4.79±0.45 | 3.89-5.75 |

Abbreviations: 5C – 5 Compartment model, D<sub>2</sub>O – deuterium dilution total body water, ISOK – isokinetic, Ext – extension, ADP BV – BodPod body volume, Db – body density
\* sex differences
† differs significantly from the value calculated if 73.2% hydration is assumed.
Note: The reference body is based on Broek's 1963 cadaver values.

**Table 2** outlines the different races, sports, and sports types, with the majority of the population being white and equal distribution of sport type. Additionally, only participants with values for each determination of FM and FFM were used for an analysis of variance test, which is described in **Figure 1** and shows no statistical mean differences between all devices of FM or FFM in either sex.

**Figure 1.**
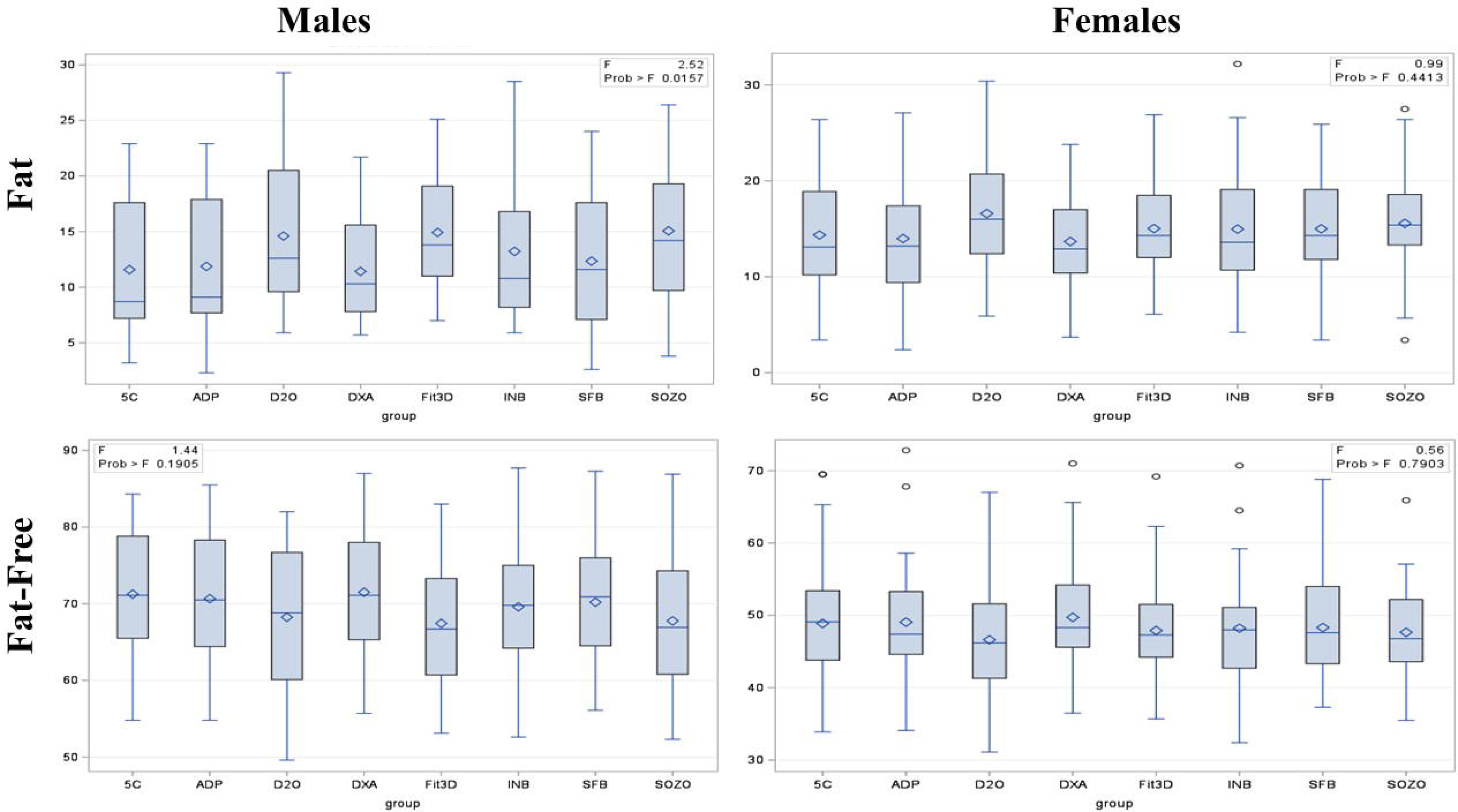
Fat and Fat-Free Mass Mean Difference in all Methods(n=75)

**Table 2.** Sport played, race, and Anaerobic / aerobic sport Distribution(n=75)

|  |  | Male | Female |
| --- | --- | --- | --- |
|  | Race | Frequency | Frequency |
|  | Asian | 11 | 6 |
|  | Black | 1 | 5 |
|  | Hispanic | 1 | 3 |
|  | NHOPI | 3 | 5 |
|  | White | 24 | 21 |
|  | Sport |  |  |
|  | Basketball | 1 | 1 |
|  | Beach Volleyball | 0 | 4 |
|  | Cheerleading | 2 | 2 |
|  | Diving | 4 | 1 |
|  | Gymnastics | 0 | 1 |
|  | MMA | 5 | 2 |
|  | Power Lifting | 5 | 2 |
|  | ROTC | 6 | 2 |
|  | Soccer | 1 | 2 |
|  | Softball | 2 | 0 |
|  | Swimming | 7 | 2 |
|  | Tennis | 0 | 1 |
|  | Track | 3 | 16 |
|  | Volleyball | 4 | 0 |
|  | Water Polo | 0 | 4 |
|  | Sport Type |  |  |
|  | Anaerobic | 17 | 11 |
|  | Aerobic | 23 | 29 |

Abbreviations: MMA – mixed martial arts NH – non-Hispanic, NHOPI – native Hawaiian or other Pacific Islander, ROTC – reserve officers' training corps Anaerobic (Without O<sub>2</sub>) = Beach Volleyball, Cheerleading, Diving, Gymnastics, Power Lifting, Softball, Tennis, Volleyball Aerobic (With O<sub>2</sub>) = Basket Ball, MMA, ROTC, Running, Soccer, Swimming, Track, Water Polo

The hydration status(TBW/FFM) of the athletes was significantly outside of the normal range(*p*=0.001) for both males(0.63-0.73%) and females(0.58-0.78%). An attempt was made to increase the accuracy of each BIA device’s TBW estimation to the criterion by using measures of skin temperature and moisture by using step-forward linear regression including each estimate of TBW along with the skin temperature and moisture variables from all locations(head, hand, and foot). However, none of the candidate variables of skin moisture or temperature were selected in the final model to increase the performance of the BIA-reported TBW values.

**Table 3** presents the results of linear regression analysis for bivariate comparisons of FM, FFM, and BV measurements using various methods, along with their agreement and precision values of each method to the criterion 5CM. The results reveal that ADP and DXA demonstrated the highest agreement in FM to the criterion(CCC=0.90-0.99), with ADP exhibiting substantial agreement(CCC=0.96) in FM for males. ADP also showed substantial agreement in FFM both sexes, while DXA and D_2_O had moderate agreement. D_2_O produced the lowest RMSE in males(1.66kg) and females(1.70kg), which was lower than ADP or DXA. The 3DO method had a moderate agreement in females FFM(CCC=0.91), the only field method to produce such an agreement in body composition. However, the 3DO had the highest precision among all methods, for both males(5.8%) and females(4.6%). Similarly, the SFB7(3.3-3.2%) and the SOZO(2.3-4.5%) had high precision estimates in males and females. All measures of FFM had excellent precision of <1.0%, whereas only ADP, DXA, D_2_O, UWW, and S10 had <2.0%. The lowest precision estimates were the S10(0.09%) in males and D2O(0.06%) in females. Ultimately, the 5CM had higher precision than most estimates(3.8-2.9% in males and females), however similar performance to previously reported criterion models ^37^. This highlights the value of this model for frequent monitoring of body composition change. All other methods(UWW and BIA did not produce high equivalence in any category(CCC<0.90).

**Table 3.** Body Composition Agreement between Methodologies to the Criterion 5 Compartment Model(n=75)

| Method | Precision |  | Male Accuracy |  |  |  |  |  | Precision |  | Female Accuracy |  |  |  |  |  |
| --- | --- | --- | --- | --- | --- | --- | --- | --- | --- | --- | --- | --- | --- | --- | --- | --- |
|  | Mean | RMS-CV | CCC | R<sup>2</sup> | RMSE | Slope | Int | Slope Int=0 | Mean | RMS-CV | CCC | R<sup>2</sup> | RMSE | Slope | Int | Slope Int=0 |
| Criterion 5C FM | 11.58 | 3.80 |  |  |  |  |  |  | 14.36 | 2.99 |  |  |  |  |  |  |
| ADP | 11.87 | 1.58 | <sup>b</sup>0.96 | 0.91 | 1.73 | 1.01 | -0.37 | 0.98 | 13.98 | 1.05 | <sup>c</sup>0.93 | 0.87 | 2.03 | 0.93 | 0.97 | 0.99 |
| D<sub>2</sub>O | 14.61 | 1.39 | 0.86 | 0.92 | 1.66 | 0.87 | -1.24 | 0.80 | 16.60 | 0.58 | 0.89 | 0.90 | 1.70 | 0.95 | -1.29 | 0.88 |
| DXA | 11.42 | 1.43 | <sup>c</sup>0.90 | 0.88 | 2.01 | 1.24 | **-2.59** | 1.04 | 13.68 | 1.26 | <sup>c</sup>0.91 | 0.87 | 2.08 | 1.08 | -0.81 | 1.03 |
| Fit3D | 14.92 | 5.82 | 0.66 | 0.64 | 3.56 | 0.98 | -3.35 | 0.78 | 15.04 | 4.57 | 0.86 | 0.78 | 2.70 | 1.02 | -1.50 | 0.93 |
| S10 | 13.22 | 0.94 | 0.72 | 0.55 | 3.95 | 0.50 | **5.37** | 0.83 | 14.97 | 1.10 | 0.80 | 0.64 | 3.40 | 0.72 | **2.78** | 0.87 |
| SFB | 12.34 | 3.33 | 0.56 | 0.28 | 4.99 | 0.61 | **4.71** | 0.86 | 15.00 | 3.19 | 0.80 | 0.64 | 3.39 | 0.86 | 1.38 | 0.95 |
| SOZO | 15.07 | 2.32 | 0.57 | 0.44 | 4.43 | 0.68 | 1.36 | 0.76 | 15.56 | 4.45 | 0.80 | 0.65 | 3.32 | 0.88 | 0.53 | 0.91 |
| UWW | 9.01 | 1.25 | 0.89 | 0.84 | 1.91 | 0.70 | 2.22 | 0.88 | 10.74 | 1.49 | 0.68 | 0.53 | 3.89 | 0.79 | 4.80 | 1.15 |
| Criterion 5C FFM | 48.87 | 0.10 |  |  |  |  |  |  | 71.25 | 0.11 |  |  |  |  |  |  |
| ADP | 49.05 | 0.07 | <sup>b</sup>0.97 | 0.95 | 1.95 | 0.96 | 3.64 | 1.00 | 70.66 | 0.07 | <sup>b</sup>0.96 | 0.93 | 2.12 | 0.93 | 3.47 | 0.99 |
| D<sub>2</sub>O | 46.64 | 0.07 | <sup>c</sup>0.92 | 0.95 | 1.84 | 0.95 | **6.28** | 1.04 | 68.22 | 0.08 | <sup>c</sup>0.94 | 0.95 | 1.74 | 0.95 | **4.21** | 1.04 |
| DXA | 49.72 | 0.05 | <sup>b</sup>0.96 | 0.92 | 2.37 | 1.01 | -0.80 | 0.99 | 71.50 | 0.10 | <sup>b</sup>0.96 | 0.93 | 2.14 | 0.98 | 0.80 | 0.98 |
| Fit3D | 47.89 | 1.02 | 0.81 | 0.82 | 3.59 | 1.02 | 2.90 | 1.06 | 67.42 | 1.02 | <sup>c</sup>0.91 | 0.85 | 3.03 | 1.02 | 0.76 | 1.03 |
| S10 | 48.22 | 0.04 | 0.86 | 0.76 | 4.22 | 0.85 | 12.43 | 1.01 | 69.59 | 0.12 | 0.88 | 0.78 | 3.70 | 0.86 | 7.87 | 1.01 |
| SFB | 48.33 | 0.04 | 0.78 | 0.62 | 5.23 | 0.84 | 12.47 | 1.01 | 70.20 | 0.34 | 0.89 | 0.81 | 3.48 | 0.94 | 3.53 | 1.00 |
| SOZO | 47.66 | 0.12 | 0.78 | 0.71 | 4.56 | 0.84 | **14.45** | 1.05 | 67.76 | 0.52 | 0.88 | 0.82 | 3.36 | 1.02 | 0.53 | 1.02 |
| UWW | 75.30 | 0.41 | 0.79 | 0.58 | 2.32 | 0.74 | 19.87 | 1.00 | 53.84 | 0.44 | 0.82 | 0.74 | 3.58 | 0.77 | 10.03 | 0.95 |
| Criterion ADP Volume | 61.10 | 0.04 |  |  |  |  |  |  | 77.42 | 0.04 |  |  |  |  |  |  |
| DXA | 49.72 | 0.03 | <sup>a</sup>0.99 | 1.00 | 0.52 | 0.01 | 0.44 | 0.98 | 78.66 | 0.04 | <sup>a</sup>0.99 | 1.00 | 0.68 | 0.01 | 0.86 | 1.03 |
| Fit3D | 58.33 | 0.03 | <sup>a</sup>0.99 | 1.00 | 0.78 | 0.01 | 0.64 | 0.98 | 76.08 | 0.08 | <sup>a</sup>0.99 | 0.99 | 1.19 | 0.02 | 1.49 | 1.02 |
| UWW | 60.89 | 0.41 | <sup>a</sup>0.99 | 1.00 | 0.86 | 1.01 | -0.49 | 1.00 | 78.48 | 0.44 | <sup>a</sup>0.99 | 0.99 | 0.86 | 0.92 | 6.30 | 1.00 |

Abbreviations: 5C – 5 Compartment model, D<sub>2</sub>O – deuterium dilution total body water.
Note: R<sup>2</sup> is adjusted, RMSE is in liters for volume measures, whereas all other measurements are in kg. Int – intercept, (Slope Int=0) is the slope, when the intercept is equal to zero. UWW was calculated on a separate sample with matched 5CM comparisons where n=24.
<sup>a</sup> = >0.99 ‘almost perfect’ equivalence
<sup>b</sup> = 0.95 to 0.99: ‘substantial’ equivalence
<sup>c</sup> = 0.90 to 0.95: ‘moderate’ equivalence
Otherwise unmarked is considered <0.90 poor equivalence
Bolded text indicates Significant Intercept

Despite a lack of a significant mean difference, **Figure 2** illustrates that devices show large individual errors and some methods did not show equivalence to the criterion, showing large 95% limits of agreement(LOA) present for each device and considerable offsets from the line of identity. Although the S10, SFB, and SOZO methods all had a CCC=0.88-0.89, indicating approaching moderate equivalence, the figure shows considerable underestimation of FM in athletes, but less so for females. Specifically, D_2_O tended to underestimate FM in almost all cases, and to a lesser extent, the 3DO device. A similar divergence was present in the FM for DXA males, where it tended to overestimate in the lower ranges and underestimate in the higher ranges.

**Figure 2.**
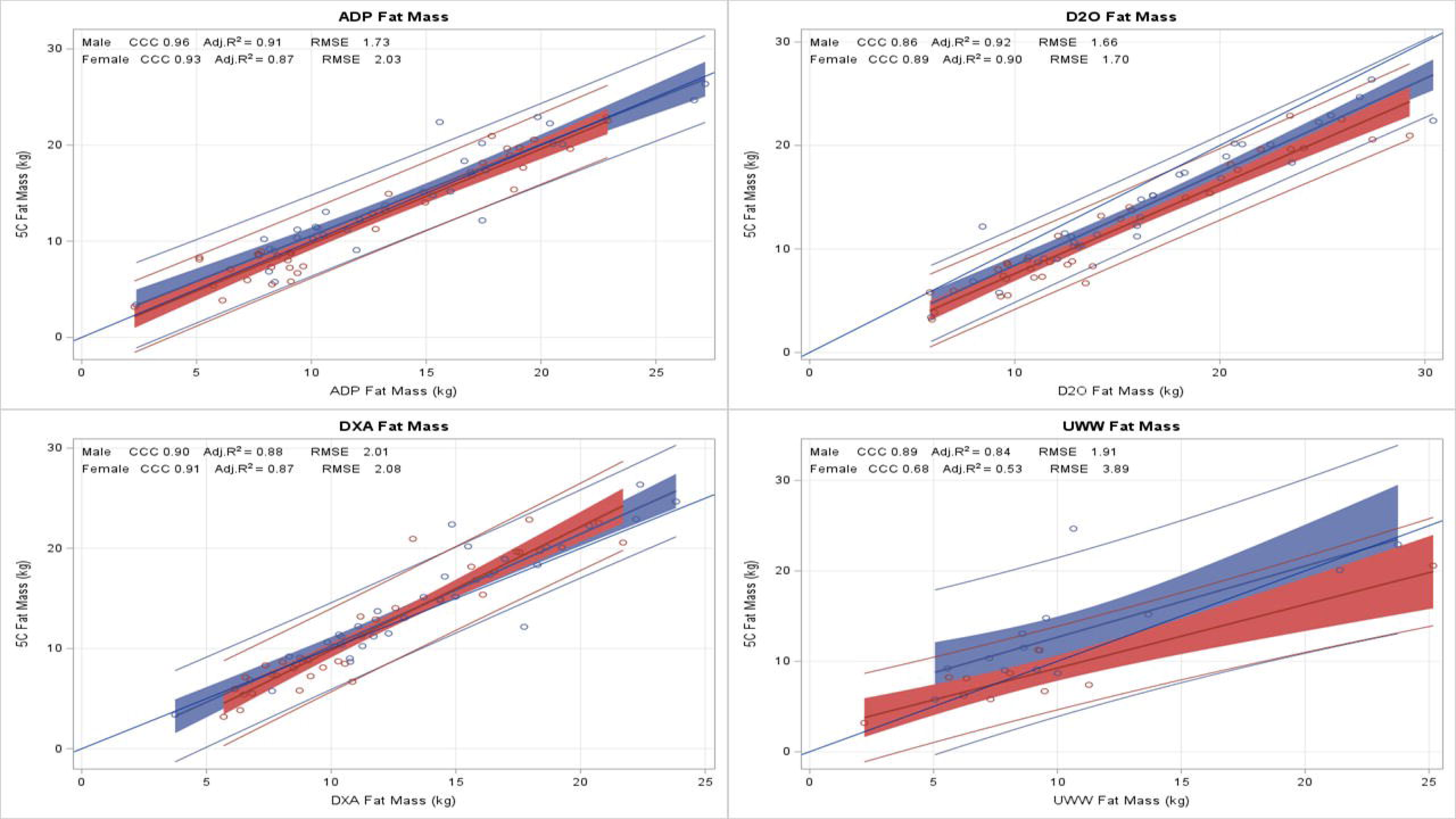
Fat Mass Agreement to the 5-Compartment Model(n=75)

Interestingly, a near-moderate agreement(CCC=0.88-0.89) was observed in FFM for the S10, SFB, and SOZO, as shown in **Figure 3**, which depicts the regression plots of each device-reported FFM to the criterion, indicating tighter ranges of 95% confidence around the line of identity. Lastly, all three volume methods using 3DO, DXA, and UWW produced nearly perfect agreement of CCC=0.99 and precision(>1%) in both males and females. However, only DXA produced high agreement and precision in body composition.

**Figure 3.**
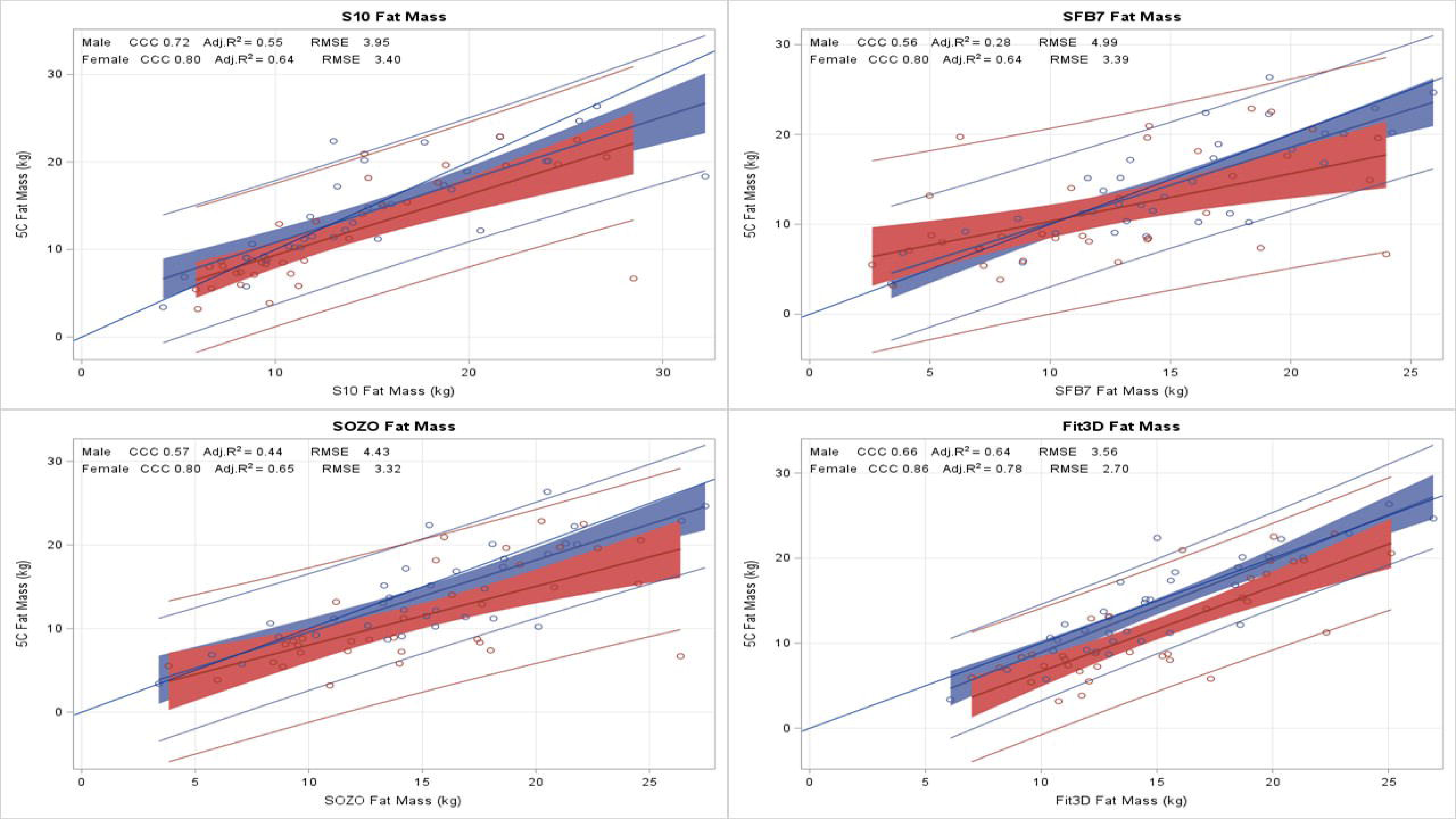
Fat-Free Mass Agreement to the 5-Compartment Model(n=75)

The estimated FFM values from each device in the determination of muscle strength via isokinetic movements of the thigh and trunk are shown in **Table 4**. Height and weight were chosen as the base model for comparison, derived using stepwise forward regression of demographic information. Although skin temperature and moisture variables were considered, they were not significant for the model. The different methods of FFM from each device produced varying estimates of muscle strength, and no single predictor of strength was significant over the other methods for males and females due to large confidence intervals and overlapping effects including the base model. However, the 5CM had the highest overall performance in each category of isokinetic leg and trunk strength for both sexes. The 5CM FFM had moderately predictive R^2^ in male and female leg strength(R^2^=0.46, 0.58, respectively), and male and female trunk strength was best predicted by the 5CM FFM(R^2^=0.55, 0.71, respectively). Devices such as ADP, DXA, and D2O all agreed the highest to the criterion 5CM for body composition measures were in the highest end of muscle strength associations, including the example of a high agreement(CCC=0.91) for 3DO in FFM to the 5CM in females also showed one of the highest associations to muscle strength(R^2^=0.69).

**Table 4.** Highest Ranking Determination of Leg and Trunk Strength by Body Composition Methods of Males and Females(n=75)

|  |  |  |  | Male |  |  |  |
| --- | --- | --- | --- | --- | --- | --- | --- |
| FFM | Leg ISOK Ex |  |  | FFM | Trunk ISOK Ex |  |  |
|  | R<sup>2</sup> | 95% CI | RMSE |  | R<sup>2</sup> | 95% CI | RMSE |
| 5C | 0.46 | 0.23-0.69 | 25.5 | 5C | 0.55 | 0.35-0.77 | 47.2 |
| D<sub>2</sub>O | 0.42 | 0.18-0.67 | 26.3 | DXA | 0.53 | 0.3-0.74 | 45.2 |
| DXA | 0.40 | 0.15-0.62 | 29.3 | S10 | 0.51 | 0.29-0.73 | 47.4 |
| ADP | 0.40 | 0.18-0.64 | 26.9 | D<sub>2</sub>O | 0.49 | 0.22-0.74 | 50.0 |
| UWW | 0.40 | 0.18-0.64 | 27.8 | ADP | 0.48 | 0.23-0.73 | 48.8 |
| SFB | 0.35 | 0.14-0.63 | 30.5 | UWW | 0.48 | 0.23-0.73 | 49.2 |
| SOZO | 0.32 | 0.12-0.61 | 30.3 | SOZO | 0.46 | 0.22-0.71 | 48.8 |
| S10 | 0.31 | 0.11-0.61 | 30.4 | Fit3D | 0.46 | 0.19-0.73 | 48.1 |
| Fit3D | 0.31 | 0.14-0.68 | 29.7 | SFB | 0.40 | 0.16-0.68 | 54.6 |
| *Base Model* | *0.25* | *0.05-0.54* | *31.3* | *Base Model* | *0.38* | *0.13-0.66* | *49.1* |

|  |  |  |  | Female |  |  |  |
| --- | --- | --- | --- | --- | --- | --- | --- |
| FFM | Leg ISOK Ex |  |  | FFM | Trunk ISOK Ex |  |  |
|  | R<sup>2</sup> | 95% CI | RMSE |  | R<sup>2</sup> | 95% CI | RMSE |
| 5C | 0.58 | 0.33-0.75 | 21.3 | 5C | 0.71 | 0.43-0.87 | 23.7 |
| D<sub>2</sub>O | 0.56 | 0.31-0.78 | 21.4 | ADP | 0.69 | 0.43-0.88 | 22.7 |
| Fit3D | 0.50 | 0.23-0.76 | 21.2 | DXA | 0.69 | 0.43-0.88 | 18.3 |
| DXA | 0.47 | 0.05-0.57 | 24.2 | Fit3D | 0.69 | 0.29-0.91 | 18.9 |
| ADP | 0.46 | 0.24-0.7 | 21.9 | D<sub>2</sub>O | 0.66 | 0.39-0.86 | 25.0 |
| UWW | 0.46 | 0.24-0.7 | 25.1 | S10 | 0.64 | 0.33-0.88 | 25.4 |
| SOZO | 0.45 | 0.19-0.72 | 22.1 | SOZO | 0.64 | 0.32-0.86 | 25.8 |
| SFB | 0.39 | 0.11-0.68 | 23.6 | SFB | 0.62 | 0.3-0.85 | 28.5 |
| S10 | 0.38 | 0.1-0.69 | 24.1 | UWW | 0.61 | 0.36-0.8 | 17.4 |
| *Base Model* | *0.18* | *0.02-0.49* | *24.4* | *Base Model* | *0.49* | *0.2-0.73* | *19.7* |

Abbreviations: 5C – 5 Compartment model, D<sub>2</sub>O – deuterium dilution total body water, FFM – fat-free mass, ADP BV – BodPod body volume, ISOK – isokinetic, Ext – extension
Note: R<sup>2</sup> is adjusted. The base model was derived using demographic information on height and weight. All RMSE are in Nm. UWW was calculated on a separate sample with matched 5C comparisons where n=24.

Furthermore, knee isometric extension/flexion and trunk/knee isokinetic flexion comparisons were conducted, and the Pearson’s correlation of each FFM estimate result is reported in **Supplemental Table 1**. Overall, females showed higher statistically significant(*p*<0.05) associations than males in all methods. The quartile *p*-trend was determined using individual FFM and their association with leg and thigh muscle strength, as represented in **Figure 4**. All FFM methods had significant *p*-trend associations(p=0.05), but some were more prominent than others, such as the 5CM in male leg strength, which had a more linear distribution over SFB7, with each column consecutively built on the next.

**Figure 4.**
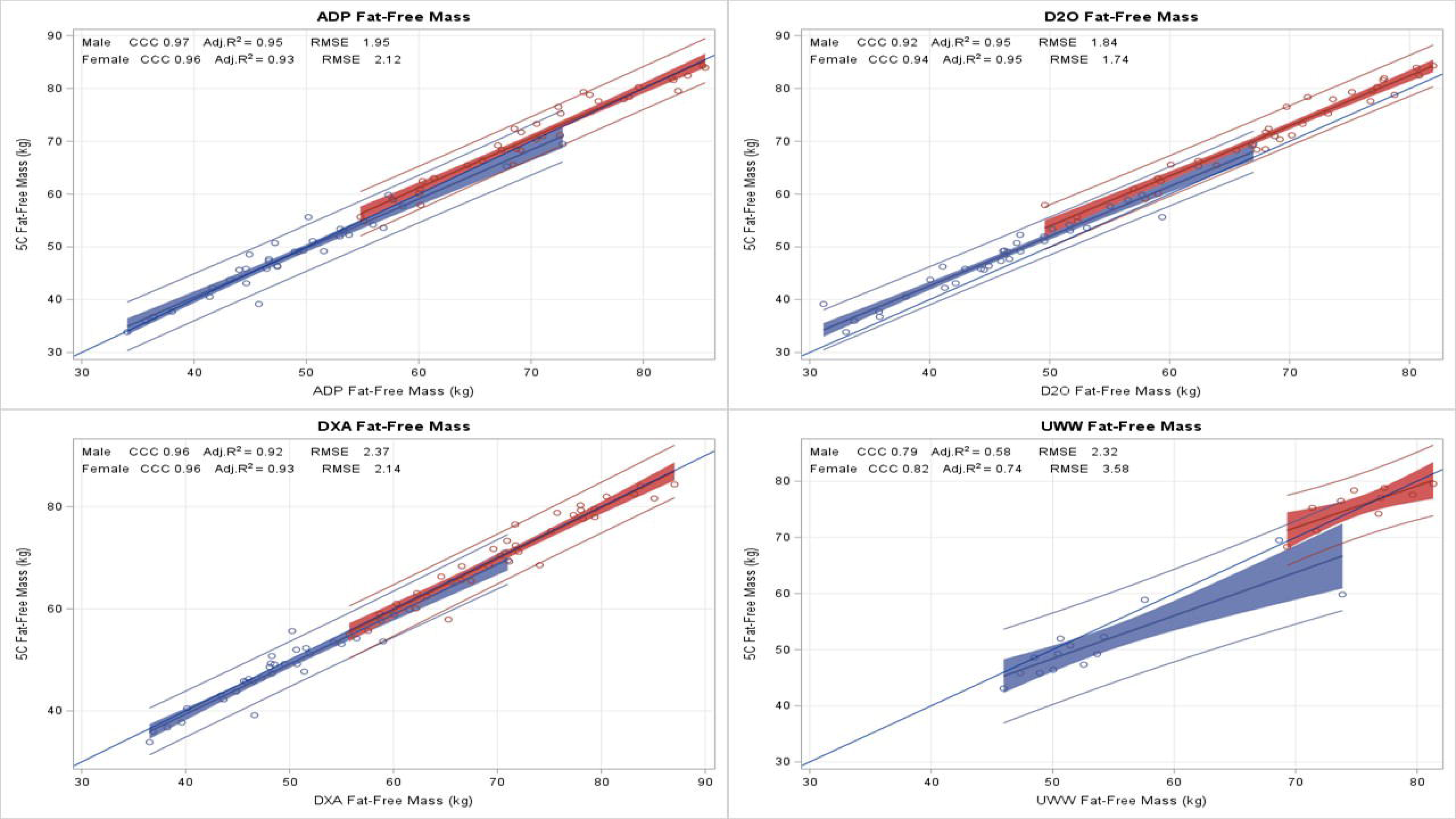
Quartile p-trend Associations of FFM Estimates Leg to Trunk Strength in Males and Females(n=75)

## DISCUSSION

This research aimed to compare various methods of body composition analysis in athletes who exhibit different levels of hydration with the criterion 5CM and evaluate their relationship with muscle strength. The findings of this study demonstrated that precise and accurate estimates of body composition can be obtained in athletes, and a more precise measurement of body composition led to better muscle strength estimates. The most reliable laboratory-based techniques were ADP and DXA, while field assessments had moderate to poor agreement. The 5CM method exhibited the strongest correlation with muscle strength among all body composition analysis techniques. Methods that showed significant agreement with body composition tended to produce a more valid determination of muscle strength in both male and female athletes. Accurate body composition estimates produce more precise muscle strength estimates in athletes, irrespective of their hydration status. ADP and DXA are trustworthy approaches for evaluating body composition and muscle strength compared to the criterion approach.

Our study contrasts with Moon, Silva, and Kendall’s research on the validity of different laboratory methods(DXA, ADP, UWW) for estimating body composition in athletes using criterion 5CM^38-40^. Unlike their separate studies, our study examined all devices together. Moon found acceptable error ranges of percent fat for all devices(except DXA), while our study found that ADP and DXA had better agreement with the line of identity in females, and UWW had the lease agreement. Silva concluded that DXA and ADP were imprecise and invalid for individual body fat prediction in athletes, but our study found moderate to substantial agreement for both sexes with DXA and ADP, and poor agreement with UWW. Kendall found poor agreement(CCC=0.84) between ADP and 5CM estimates for FFM in male athletes, while our study found substantial agreement(CCC=0.97) for ADP. All researchers agree that a multicompartment model criterion(4/5C) provides the most accurate assessment of body composition in athletes, considering the wide range of hydration values found in our study. The remaining devices, such as BIA and 3DO, had larger errors at the individual level, possibly due to methodological assumptions that the criterion methods lack. Hence, further advancements are necessary to improve their accuracy and clinical relevance. Our volume estimates align with previous findings, except for the 3DO showing better precision than reported before^41^. Other studies have also successfully utilized 3DO images for reliable predictions of body composition^42^.

Our study agrees with other studies that estimate muscle strength using FFM such as Buehring, Raymond-Pope, and Bourgeois^43-45^. However, considerable sex differences related to body size are relevant when predicting muscle strength with body composition. In our study, DXA FFM was associated with leg strength with R^2^=0.40 males and R^2^=0.47 females. Non-significant relationships, or those with relatively low correlation, may become significant when subgroups like sex are combined^46^. The combined association for males and females for whole body FFM and leg strength was R^2^=0.64, virtually identical to Bourgeois. We recommend reporting sex-specific muscle strength associations to avoid this type of correlation inflation and to not generalize relationships across sexes.

### Clinical Implications

The estimation of FM using BMI is inappropriate in heavily muscled males and females. Certain equations to estimate body composition using BMI work reasonably well for a population of normal aerobic fitness males and females, but generally, BMI cannot distinguish the high FFM of FM component of individuals who exceeded a BMI of 30 kg/m^2^. Athletes who differ in height, weight, or body composition will oftentimes be incorrectly categorized by BMI class of being ‘underweight’, ‘healthy’, ‘overweight’, or ‘obese’. This is highlighted in **Figure 5** using 3D images from the current study comparing sex-based differences in five athletes with similar body FM ranges(males=9-10kg, females=19-20kg) but varies in each BMI category. The predictive values of BMI for estimating FM were poor in both males and females(R^2^=0.42, 0.41, respectively), indicating a low level of accuracy in using BMI as a predictor of FM in both sexes. Furthermore, BMI is not a better determinant of muscle strength than FFM. On the contrary, those in the ∼25 BMI range had the highest strength results over other BMI categories. Potentially suggesting that an increase in FM may hinder strength performance.

**Figure 5.**
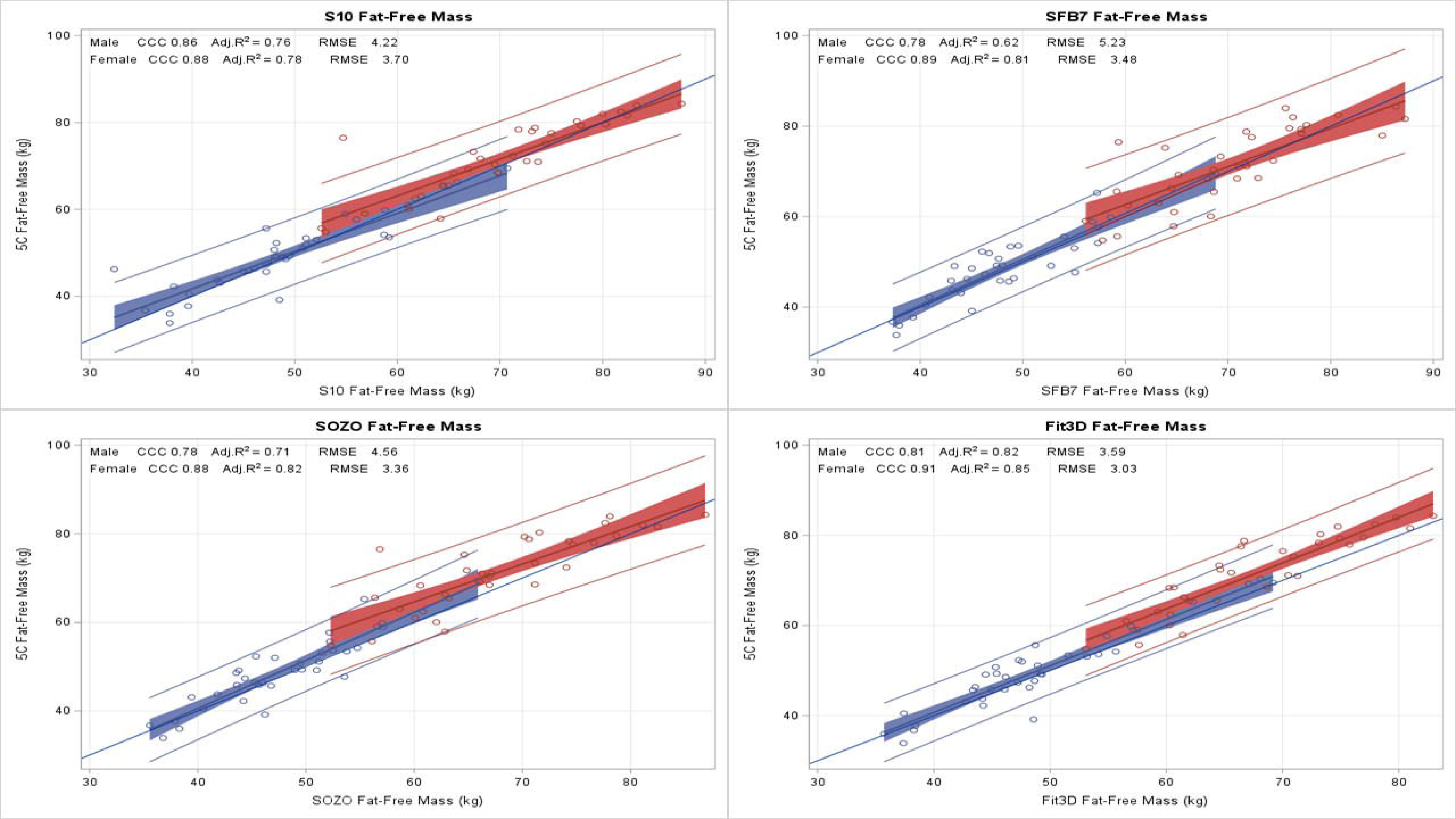
A Comparative Analysis of Sex-Based Differences in Fat Mass and BMI among Five Athletes with a Fat Mass Range of 9 – 10kg for Males and 19-20kg for Females

To our knowledge, this is the first investigation evaluating the body composition agreement between criterion 5CM and other laboratory and field methods, specifically the associations of each device-reported FFM estimate to muscle strength in a collegiate athletic population. Another strength of our study is that the criterion of muscle strength was measured in large muscle groups like the legs and trunk in multiple movements of isokinetic and isometric. Lastly, both males and females were separated in this analysis to more accurately report expected correlations by sex.

### Limitations

Although we feel that measurement of the thigh and abdominal/back muscles are more functionally relevant than grip strength, we were unable to directly compare our results to much of the muscle strength literature. Due to the small sample size, we were unable to withhold a subset of participants for model testing. Increasing the sample size should improve our ability to discern statistically significant differences by technique to strength.

### Conclusion

From this investigation, we conclude that, when assessing body composition and estimates of muscle strength, researchers and clinicians should evaluate which device is to be used based on its accuracy in comparison to a criterion method, such as the 5CM. This is due to the results demonstrating that the more advanced methods of body composition analysis tend to demonstrate higher agreement and thus support a stronger association with muscle strength than the more commonly used methods. Furthermore, the 5CM is particularly effective in estimating isokinetic and isometric muscle strength, further compounding its utility. Future research in athletes should examine the effects of changes in FFM due to training, weight loss, and/or gain on functional measures when compared to a criterion method.

## Data Availability

All data produced in the present study are available upon reasonable request to the authors

https://shepherdresearchlab.org/

## Takeaways

1. Although the 5-compartment model is the most accurate estimate of *in vivo* body composition, it is not practical for clinicians and coaches outside of laboratory settings. More accessible assessment methods should provide reasonable accuracy to the criterion.
2. Commercially-available accessible methods produced considerable differences in fat mass and fat-free mass compared to the criterion estimates ranging from substantial to poor agreement to the criterion in both sexes. Air-displacement plethysmography and DXA produced the highest agreement in all categories.
3. Accessible methods that showed substantial concordance(agreement) to 5-compartment body composition tended to produce a more valid model of muscle strength in both males and females of a cohort of athletes.

*Non-finding*: Skin temperature and moisture were not significant in improving body composition or strength predictors.

### What is already known on this topic

Body composition and muscle strength are significant predictors of athletic performance.

### What this study adds

An ideal body composition assessment approach would provide valid estimates to the criterion and strongly link to a functional component such as muscle strength. This study provides validations of assessment methods for fat and fat-free mass and evaluates their associations with isokinetic and isometric muscle strength.

### How this study might affect research, practice, or policy

This investigation provides clinicians and coaches with information vital to identifying the optimal tool for monitoring body composition and strength in laboratory and field settings.

## Abbreviations

3DO: 3-dimensional optical
BIA: bioelectrical impedance analysis
D2O: deuterium
TBW: total body water
DXA: dual-energy X-ray absorptiometry
BMC: bone mineral content
ADP: air displacement plethysmography
BV: body volume
BM: body mass
Mo: osseous mineral
Ms: soft tissue mineral
FFM: fat-free mass
FM: fat mass
kg: kilogram
L: liter
NHOPI: Native Hawaiian or Pacific Islander
NH: non-Hispanic
SD: standard deviation
CCC: Lin’s concordance correlation coefficient
RMSE: root mean square error, root-mean-square coefficient of variation(RMS-CV%)

## Acknowledgment

We gratefully acknowledge En Liu and Nisa Kelly for subject recruitment and implementation of the study protocol. We would also like to acknowledge Timothy Shriver(University of Wisconsin), and Valery Hymel(Pennington Biomedical Research Laboratory) for processing and analyzing all D_2_O specimens. This study was funded by Hologic Inc. grant # 2018-01102.

## Author Contributions

John Shepherd, Steven B. Heymsfield, and Thomas Kelly: Conceptualization, Funding acquisition Brandon Quon and Devon Cataldi: Data curation, Formal analysis John A Shepherd: Investigation, Validation, Visualizations Devon Cataldi, Jonathan Bennett, Michael Wong, Nisa Kelly, and Yong En Liu: Methodology, Project administration, Resources, Software, Supervision Devon Cataldi, John Shepherd, Steven B. Heymsfield, Dale A. Schoeller and Jonathan Bennett: Roles/Writing -Original draft, review & editing Data Share Statement: Data described in the manuscript, codebook, and analytic code will be made available upon request pending an application and approval, payment, and or other.

## Supplemental

*Supplemental Table 1 – Description of Body Composition Methods*

*Supplemental Table 2 – Pearson’s Correlation of Differing Body Composition Methods to Leg and Trunk Isokinetic and Isometric, Flexion and Extension Strength(n=75)*

**Supplemental Figure 1.**
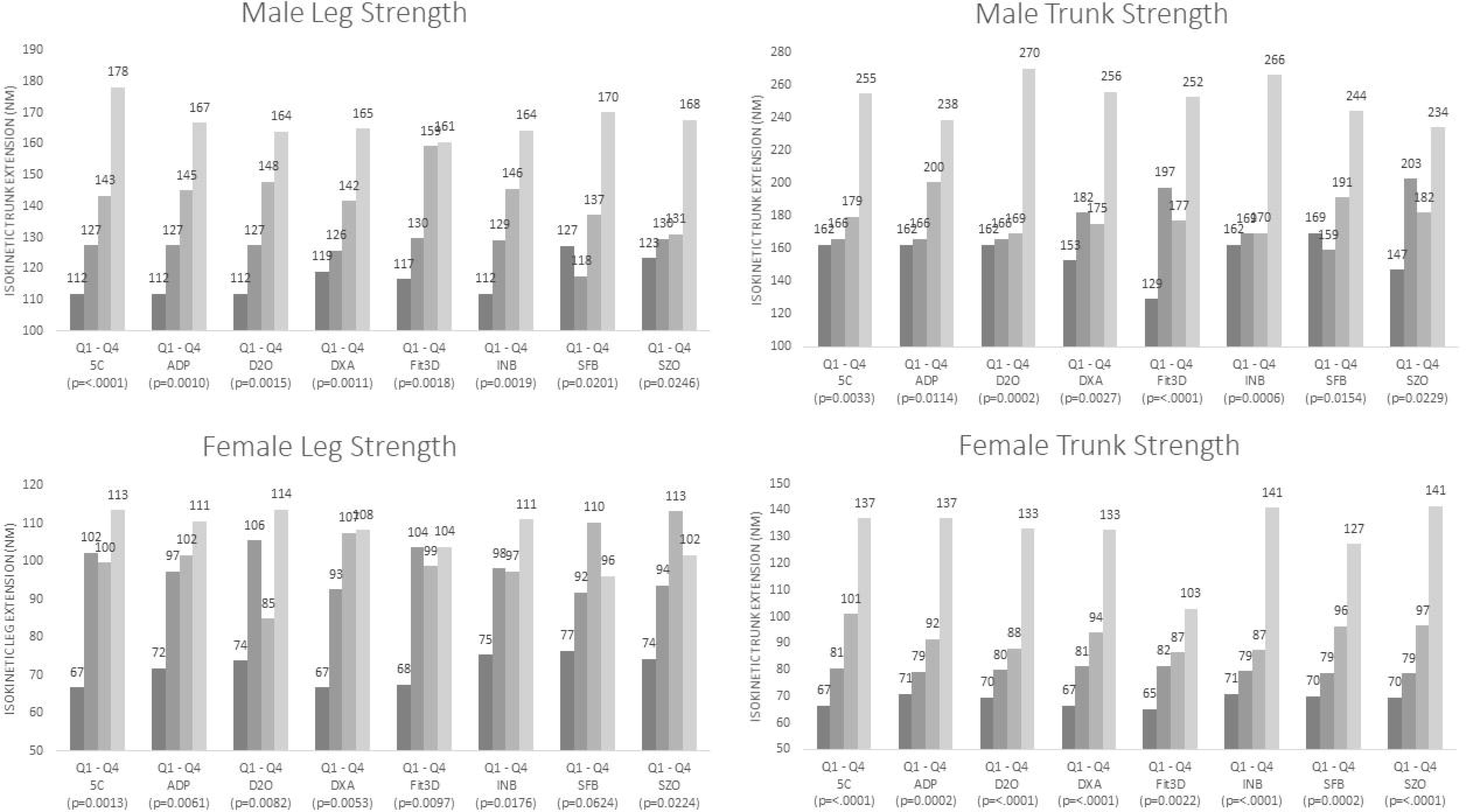
Consort Statement

