## Supplemental for "Accuracy and Precision of Multiple Laboratory and Field Methods to The Criterion *In Vivo* Five-Compartment Body Composition Model and Their Association with Muscle Strength in Collegiate Athletes of Varying States of Hydration: The *Da Kine* Protocol Study"

Supplemental Table 1 – Description of Body Composition Methods

| Method | Assumptions | Advantages | Disadvantages |
| --- | --- | --- | --- |
| 5CM | The criterion for body composition | Criterion Method for fat mass | Not clinically practical. Propagation of measurement error |
| DXA | Constant Densities | Quick, provides segmental body composition | Expensive, time-consuming, and low radiation exposure |
| D_2_O | Isotopic tracer method errors | Criterion method for total body water | Expensive, time-consuming |
| ADP | Constant Densities | Criterion Method for body volume | Expensive, time-consuming |
| UWW | Constant Densities | Criterion method for body volume | Burdensome on the participant, time-intensive |
| BIA | Population Specific | Quick, inexpensive | Not accurate in all participants |
| 3DO | Device Specific | Quick, cheap, and accurate for body volume | Accuracy for body composition is questionable |

Abbreviations: Anthro–Anthropometry, 5C – 5 Compartment model, D_2_O – deuterium dilution total body water

*Supplemental Table 2 – Pearson’s Correlation of Differing Body Composition Methods to Leg and Trunk Isokinetic and Isometric, Flexion and Extension Strength (n=75)*

|  | Male | | | | | | Female | | | | | |
| --- | --- | --- | --- | --- | --- | --- | --- | --- | --- | --- | --- | --- |
|  | ISOK LEG Ext | ISOK LEG FLEX | ISOM LEG Ext | ISOM LEG FLEX | ISOK TRK Ext | ISOK TRK FLEX | ISOK LEG Ext | ISOK LEG FLEX | ISOM LEG Ext | ISOM LEG FLEX | ISOK TRK Ext | ISOK TRK FLEX |
| Height | **0.47** | 0.30 | 0.30 | 0.31 | **0.59** | 0.16 | **0.56** | **0.48** | **0.50** | **0.49** | **0.77** | **0.67** |
| Weight | 0.27 | 0.25 | 0.30 | 0.26 | **0.42** | 0.08 | **0.47** | **0.54** | **0.51** | **0.53** | **0.63** | **0.58** |
| 5C | **0.64** | **0.41** | **0.43** | **0.42** | **0.69** | 0.25 | **0.71** | **0.67** | **0.65** | **0.71** | **0.81** | **0.73** |
| ADP | **0.58** | **0.37** | **0.38** | **0.37** | **0.66** | 0.20 | **0.66** | **0.67** | **0.61** | **0.72** | **0.82** | **0.74** |
| D_2_O | **0.60** | **0.44** | **0.38** | **0.41** | **0.63** | 0.25 | **0.72** | **0.65** | **0.64** | **0.69** | **0.78** | **0.70** |
| DXA | **0.56** | **0.37** | **0.38** | **0.40** | **0.71** | 0.24 | **0.65** | **0.62** | **0.60** | **0.67** | **0.83** | **0.76** |
| Fit3D | **0.53** | **0.33** | **0.36** | 0.31 | **0.63** | 0.09 | **0.60** | **0.57** | **0.57** | **0.63** | **0.82** | **0.73** |
| S10 | **0.44** | **0.34** | **0.36** | **0.36** | **0.68** | 0.22 | **0.54** | **0.56** | **0.46** | **0.65** | **0.76** | **0.72** |
| SFB | **0.50** | 0.31 | **0.37** | 0.22 | **0.49** | 0.11 | **0.55** | **0.48** | **0.51** | **0.61** | **0.69** | **0.64** |
| SOZO | **0.50** | **0.32** | **0.33** | 0.28 | **0.63** | 0.25 | **0.62** | **0.52** | **0.56** | **0.60** | **0.75** | **0.69** |
| UWW* | **0.67** | 0.60 | **0.64** | 0.54 | **0.70** | 0.66 | **0.68** | 0.54 | **0.82** | **0.60** | **0.92** | **0.77** |

Bold text indicates *p*<0.05.

Abbreviations: Abbreviations: 5C – 5 Compartment model, D_2_O – deuterium dilution total body water, ISOK – isokinetic, ISOM – isometric, Ext – extension, FLEX – flexion

Note: *n=24

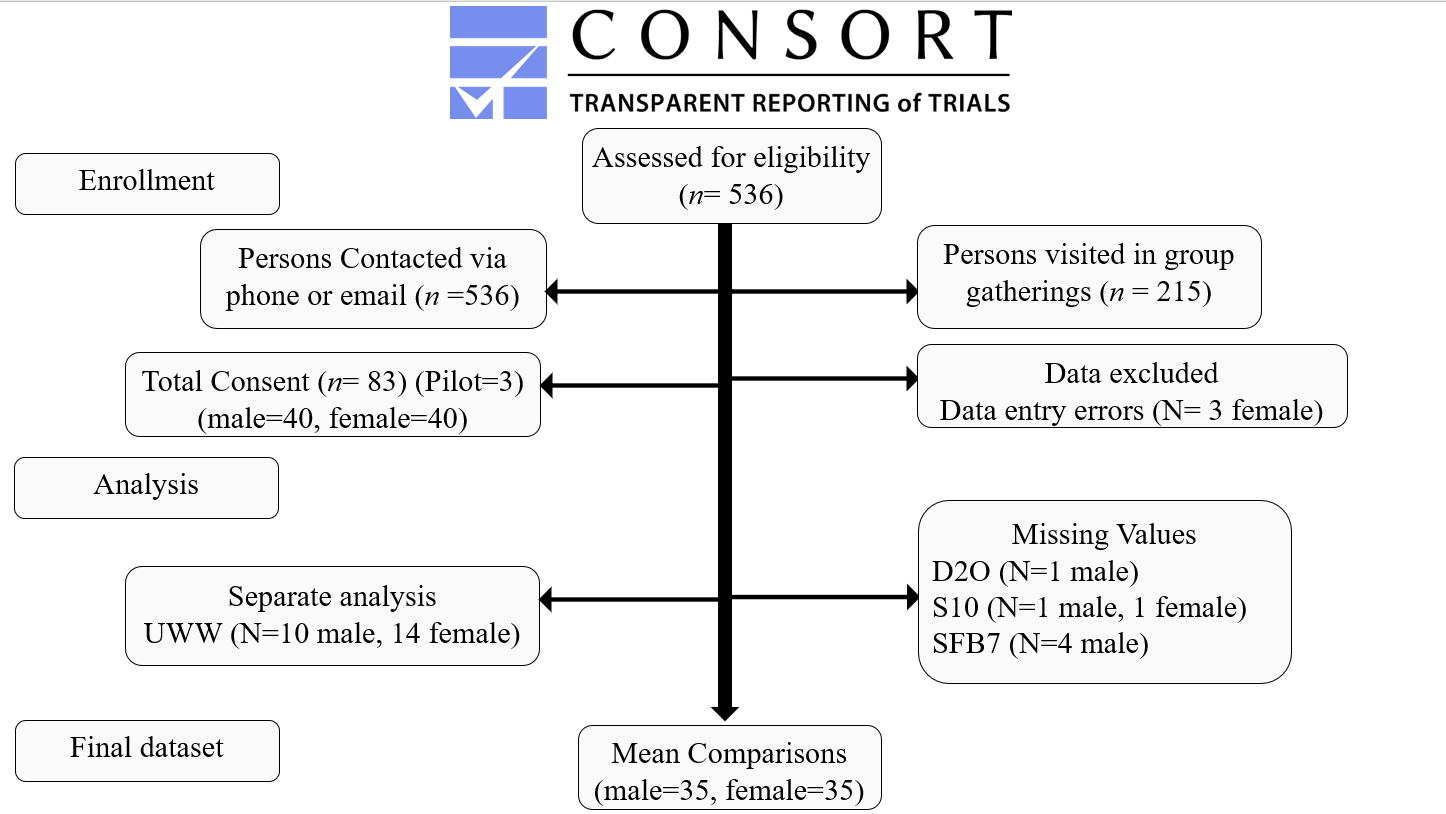

*Supplemental Figure 1 – Consort Statement*
